# Gamified interventions to improve the knowledge, awareness and practice on rational use of antibiotics among school children in Mysuru, South India to curb the growing antimicrobial resistance (AMR)

**DOI:** 10.1101/2024.04.14.24305781

**Authors:** Sumana M Neelambike, Supreeta R Shettar, Yogeesh Maheshwarappa, Praveen Kulkarni

## Abstract

**Introduction:** Antimicrobial resistance (AMR) is a global problem. It’s important to create awareness of the rational use of antibiotics to curb AMR.

**Objective:** To improve the awareness of the rational use of antibiotics through innovative games for High School Students.

**Design:** Prospective interventional study

**Setting:** Twelve High Schools in the city of Mysuru, South India.

**Participants:** 2195 Students between 13 and 16 years.

**Intervention:** Innovative gamified interventions to educate on the rational use of antibiotics. Use of antibiotics only in bacterial infections of the respiratory tract, gut, urinary tract and skin was introduced through black blackboard. An animated video was shown on the effects of irrational antibiotic use. Situations in which antibiotics can be avoided in respiratory infections and gastroenteritis were taught through Bucketing the Ball and Monkeying with Donkey games. Pre-test and Post-test questionnaires were administered and evaluated.

**Main outcome measures:** To assess the improvement in awareness of the rational use of antibiotics.

**Results:** After the intervention, knowledge of the use of antibiotics only for bacterial infections improved from 11.5% to 82.5% and ill-effects of antibiotics improved from 2.5% to 82.5%. Awareness of when not to use and when to use antibiotics for respiratory infections and gastroenteritis improved from 5.1% to 96.77%, awareness of better use of antibiotics for urinary and skin infections improved from 19.6% to 90.38% and not buying antibiotics directly from the pharmacy without prescription, and completing the antibiotic course improved from 20.3% to 91.92%. p-value < 0.05 for all components.

**Conclusions:** Awareness of the rational use of antibiotics is very poor among the general public. The need of the hour is to create awareness not only among healthcare providers but also among the general public. Innovative gamified interventions create better and long-lasting awareness of this.

**Strength and Limitations of the study:**

- The strength was using gamified interventions to create knowledge and awareness about rational use of antibiotics.
- The limitation is, awareness about adverse effects of antibiotics could not be converted to gamified intervention, and it was shown as an animated video since adverse effects of antibiotics include long term complications.
- Though the students may not be able to remember in the long run the difference between viral and bacterial infections, they will definitely be able to remember that not all respiratory tract infections and gastro enteritis require antibiotics and henceforth they will not pressurize the physicians to prescribe antibiotics.

**Limitation:** 

## Introduction

Antimicrobial resistance (AMR) is one of the most alarming problems and the toughest challenge posed to the human health this century (1–3). As the AMR crisis has hit the peak, experts have declared that the Era of ineffective antibiotics is fast approaching (4). It has been estimated by World Health Organization (WHO) that about half of antimicrobial medicines are inappropriately prescribed and about half of the patients fail to take their medicines completely(5). The clinical efficacy of antibiotics depends greatly on their correct use which in turn depends on the patients, physicians and the pharmacists (5). The lack of knowledge, widespread problems in attitudes, beliefs and behaviors has been reported among consumers which directly influences the rational antibiotic usage (5). Alongside, the physicians’ decision is influenced by several factors such as the fear of losing the patients’ trust, a lack of correct information on indications for antibiotic use and pressure from patients and family. In such cases, the patients’ expectations influence antibiotic prescription and antibiotics are more likely to be prescribed in pressured context (5). In a survey conducted by Minen et al., in north east of the United States indicated that there is need for both education and feedback on antimicrobial prescribing(6). In a study conducted by Cebotarenco and Bush, it was observed that mothers often influence medical decisions on antibiotic prescription for their children (7).In a study conducted by Tang et al., in china, it was found that among all the positive cases for Upper Respiratory tract infections (URTIs), 81.7 % were viral infections, only 11.6% of cases were bacterial and 6.7% cases were bacterial and viral co-infections (8). However, a lack of judicious differentiation of viral and bacterial causes of infections based on clinical features and lack of rapid and accurate tests for differentiation of the two has contributed to abuse of antibiotics around the world(8). Despite evidences of no benefit, misuse of antibiotics in treatment of Acute Respiratory Tract Infections (ARTIs) has been the leading cause of antibiotic prescription at outpatient visit with over 80% receiving antibiotic unnecessarily in Lower Middle Income Countries (LMICs) (9). In a study conducted by Collins et al in the United States for the period of 10 years involving patients diagnosed with viral and bacterial gastro intestinal infections from National Ambulatory Medical care Survey and National Hospital Ambulatory Medical Care Survey (NAMCS/NHAMCS), in 12.3% of viral gastro enteritis antibiotics were unnecessarily used (10).

Some of the other common infections encountered on out-patient basis are Urinary tract infections and suppurative/necrotic skin and soft tissue infections. These are most often caused by bacteria and require antibiotics for treatment (11–13). So in the community seeking out-patient health care, the respiratory tract infections and gastroenteritis are more often irrationally treated with antibiotics.

Several countries have developed campaigns to modify public misconceptions regarding the effectiveness of antibiotics, to promote appropriate use of antibiotics and prevent the development of antibiotic resistance (11, 14, and 15). In India, National Action Plan (NAP) for controlling AMR was released in April 2017 by the Indian Ministry of Health and Family Welfare (16). There are different priorities outlined in the NAP for antimicrobial resistance in India. The current NAP is comprehensive and aligns well with the World Health Organization (WHO)’s Global action plan (GAP) for AMR. The first strategy is to improve awareness and understanding of AMR through effective communication, education, and training (17). So an organizational or healthcare-system-wide approach for supporting and monitoring the prudent use of antibiotics to preserve their effectiveness is necessary among the general public, physicians and pharmacist/drug dispensers (17).

The public need to be educated on the ill-effects of unnecessary antibiotic use like entering into post antibiotic era, increased mortality and morbidity due to drug resistant infections, obesity with consequences, allergic disorders and inflammatory bowel diseases (13,18). They need to be educated that the common respiratory infections and gastroenteritis are most often caused by viruses and few by bacteria. They need to understand that most such infections rarely require antibiotic therapy. Other common infections like urinary tract infections and skin and soft tissue infections are most often caused by bacteria and thus require antibiotic therapy.

This prospective interventional study was hence taken up to create awareness among the high school students. This knowledge could help prevent unnecessary use of antibiotics and also in restraining from:

1. Using unused antibiotics available at home
2. Buying and using the antibiotics directly from pharmacy without prescription
3. Pressurising physicians to prescribe antibiotics
4. Not completing the full course of antibiotics.

## Material and methods

### Materials

High school students from 12 different JSS High Schools both English and Kannada medium, located in the most thickly populated areas of city of Mysuru, Karnataka, India mostly belonging to middle and lower socio-economic strata constituted the study material. Informed consent from each school was taken prior to the scientific awareness and intervention program. Figure 1 depicts the areas from which different schools were chosen to create awareness and table 1 depicts the identification numbers allotted to each school.

**Table 1.** Depicts the identification numbers allotted to each school.

| School Name | Numbers allotted to each school | No of Students |
| --- | --- | --- |
| JSS High School, Bannimantap | 1 | 200 |
| JSS High School, G.B.Palya | 2 | 120 |
| JSS High School, Vijaynagar | 3 | 90 |
| JSS High School, Srirampura | 4 | 120 |
| JSS High School, Lakshmipuram | 5 | 150 |
| JSS High School, Baljagath | 6 | 150 |
| JSS High School, JSS Layout | 7 | 80 |
| JSS High School, Saraswathipuram | 8 | 500 |
| JSS High School, Udaygiri | 9 | 140 |
| JSS High School, Metgalli | 10 | 200 |
| JSS High School, Nachinhallipalya | 11 | 200 |
| JSS High School, Siddharth Nagar | 12 | 245 |
| **Total** |  | **2195** |

### Methods

A total of 2195 students were included in the study. These students belonged to twelve JSS High Schools spread over the different areas of city of Mysuru. All the students were made to assemble in the auditorium. The students were divided into fifteen groups in each school. The total number of groups were 180 across 12 schools. The pre-test questionnaire was first administered to each group over a period of 15 minutes. The intervention was then conducted through the following:

1. Introduction to four types of microbes (bacteria, virus, fungi and parasite) and use of antibiotics only for bacterial infections of respiratory tract, gut, urinary tract and skin through blackboard teaching. As most of the urinary tract infections and skin and soft tissue infections are caused by bacteria, awareness was created not to neglect them and seek medical care to use antibiotics wherever necessary.
2. Animated video on ill-effects of unnecessary use of antibiotics.
3. Games on when to use and when not to use antibiotics to prevent irrational use of antibiotics.

Post-Test Questionnaire was administered to assess the effectiveness of intervention/education in all the schools (Total number of groups in 12 schools=180)

### Questionnaire

A questionnaire was designed in the local language Kannada for 30 marks that were divided into three parts. The first part consisted of two questions focusing on the appropriate use of antibiotics and the ill effects of antibiotics, where the students had to write True/False and mention any four ill effects of antibiotics respectively. The second part involved a host of features of upper and lower respiratory tract infections and gastroenteritis caused by both viruses and bacteria (5, 11, 19, and 20). The students had to tick only the bacterial symptoms which required antibiotic therapy and not the viral symptoms. The third part comprised of 10 statements, which mainly focused on the common infections that most often required antibiotics and the ill effects of antibiotics like obesity. The students had to just write true/false about these statements.

### Game Design

#### 1. Bucketing the ball (For Respiratory infections)

##### Learning Objective

Helps in understanding that not all respiratory infections require to be treated with antibiotics so that unnecessary use of antibiotics can be prevented and wherever necessary antibiotics should be administered.

##### Method

Each feature of both viral and bacterial respiratory tract infections was written on individual plastic balls. These balls were distributed to a set of students. As they played the game, each feature written on the ball was assigned to either viral or bacterial cause by the instructors. Accordingly, students were instructed to throw the ball into the two different buckets labelled as 1. *No antibiotics required* (for viral infections) and 2. *Antibiotics required* (for bacterial infections). Looking at them the other students learnt to differentiate the features of viral and bacterial infections. The next set of students who were made to play had to differentiate the different features by themselves and throw the ball from a distance into the appropriate buckets. Those students who identified the features correctly and appropriately bucketed the ball were rewarded with a pencil. Figure-2 depicts the illustration for Bucketing the ball game. Even if the students cannot differentiate the features correctly, they at least can understand that not all respiratory infections require antibiotics and consulting the doctor is important before self-medicating themselves with antibiotics irrationally.

#### 2. Monkeying with Donkey (For Gastro enteritis)

##### Learning Objective

The players get to understand who needs to be treated with antibiotics and who need not be for gastro enteritis and consult doctors before self-medication.

##### Method

Waste plastic water bottles were labelled with different features of gastroenteritis in different age group with different health conditions. The above bottles were scattered in four different boxes drawn on the floor as depicted in the figure 3. These bottles had to be sorted out by the players into two boxes one labelled -*No antibiotics necessary* and the other labelled-*Antibiotics necessary* by dodging the denner (monkeying with the denner) who walks in the fixed path as depicted in figure 3. If any of the players cannot dodge the touch of the denner, that player replaces the position of the denner and the denner gets into the game. When the players sort out the bottles appropriately dodging the denner, the denner gets the letter ‘D’ from the word Donkey. Similarly, another group plays this game and after successful completion of the game the denner gets the letter ‘O’. This game is completed sequentially by different groups till the different denners get the letters N, K, E and Y. Who so ever gets the last letter Y gets the fun tag “Donkey”.

### Outcome of the study

These three interventions created

1. Knowledge on what antibiotic is and for what type of infections antibiotics are to be used.
2. Awareness on
  a. Not using the antibiotics irrationally by self-medication
  b. Practice of not buying antibiotics directly from pharmacy without doctors’ prescription
  c. Not pressurizing the doctors for antibiotic prescriptions and
  d. Completing the course of antibiotics without stopping them in the middle.

Pre-test and Post-test questionnaires were evaluated and tabulated for further analysis on the effectiveness of the intervention/education. The results were graded as follows taking into consideration the average score of each school; Very poor for 0-5, poor for >5-10, moderate for >10-15, fair for >15-20, good for >20-25 and excellent for >25-30. The results were analysed statistically using T-test.

### Patient and Public involvement

No patients were involved in the study.

## Results

The awareness on different components assessed pre-test and post-test on rational use of antibiotics were:

I. Knowledge on use of antibiotics only in bacterial infections
II. Knowledge on ill-effects of unnecessary antibiotic use.
III. Awareness on when not to use and when to use antibiotics in common infections like respiratory tract infections and gastroenteritis.
IV. Awareness on improved use of antibiotics in urinary tract infections and skin and soft tissue infections without neglecting them as most these infections are caused by bacteria.
V. The practice of not buying antibiotics without prescription directly from pharmacy and completing the course of antibiotic prescribed by the doctors. Table 2 depicts the five different components assessed pre and post-test.

Pre-Test result analysis: The knowledge on ill-effects of antibiotics was the least during the pre-test (2.5%). The knowledge of the use of antibiotics only for bacterial infections was also very low (11.5%). The awareness of when not to use and when to use antibiotics for common infections like respiratory tract infections and gastroenteritis was also very low (5.1%). The awareness of the use of antibiotics for urinary tract infection and skin and soft tissue infection was better compared to the first three components (19.6%) in the pre-test. The practice of not buying antibiotics from the pharmacy, and completing the antibiotic course was also better compared to the first three components (20.3%) in the pre-test.

**Table-2.** Depicts the five different components assessed pre and post-test.

| **Component** | **Pre-Test in % (Out of 180 groups involving 2195 students)** | **Post-Test in % ( Out of 180 groups involving 2195 students)** | **p value** |
| --- | --- | --- | --- |
| I | 11.5 | 82.5 | 0.03 |
| II | 2.5 | 82.5 | 0.03 |
| III | 5.1 | 96.77 | 0.03 |
| IV | 19.6 | 90.38 | 0.03 |
| V | 20.3 | 91.92 | 0.03 |

After the intervention, the knowledge on ill-effects of antibiotics improved from 2.5% to 82.5% (p value < 0.05). The knowledge on use of antibiotics only for bacterial infections improved from 11.5% to 82.5% (p value < 0.05). The awareness on when not to use and when to use antibiotics for common infections like respiratory tract infections and gastro enteritis improved from 5.1% to 96.77% (p value < 0.05). The awareness on increased use of antibiotics for urinary infections, skin and soft tissue infections and not neglecting them, improved from 19.6% to 90.38% (p value < 0.05). The practice of not buying antibiotics directly from pharmacy without prescription, and completing the antibiotic course improved from 20.3% to 91.92 % (p value < 0.05). Our intervention was effective and statistically significant as all the p values are below 0.05.

As per the grading described in the methodology, in the pre-test majority of the schools had a poor knowledge and awareness (10/12 schools) and two schools had very poor performance. In the post test majority of the schools had good performance (9/12 schools), two schools had fair performance (2/12) and only one school had an excellent performance. Table-3 and figure-4 depicts the performance of different schools before and after the intervention.

**Table-3.** Depicts the performance of different schools before and after the intervention.

| **School Name** | **Pre-test -Mean ± SD and (Grading )** | **Post-test -Mean ± SD and (Grading)** | **p value** |
| --- | --- | --- | --- |
| **School-01** | 6.93 ±2.15 (Poor) | 21.73 ±3.34 (Good) | 0.001 |
| **School-02** | 5.46 ±3.11 (Poor) | 20.66 ±4.43 (Good) | 0.001 |
| **School-03** | 7.26 ±2.49 (Poor) | 22.4 ±4.45 (Good) | 0.001 |
| **School-04** | 7 ±2.2 (Poor) | 20 ±4.17 (Fair) | 0.001 |
| **School-05** | 4.33 ±2.87 (Very Poor) | 21.86 ±3.94 (Good) | 0.001 |
| **School-06** | 6.66 ±2.35 (Poor) | 26.46 ±3.39( Excellent) | 0.001 |
| **School-07** | 4.4 ± 3.04 (Very Poor) | 23.8 ± 3.52 (Good) | 0.001 |
| **School-08** | 5.53 ± 3.52 (Poor) | 19.73 ± 3.78 (Fair) | 0.001 |
| **School-09** | 6.86 ± 2.61 (Poor) | 23.46 ± 3.15 (Good) | 0.001 |
| **School-10** | 6.66 ± 2.41 (Poor ) | 24.2 ± 4.07 (Good) | 0.001 |
| **School-11** | 6.26 ± 2.71 (Poor) | 22.6 ± 4.03 (Good) | 0.001 |
| **School-12** | 6.13 ± 2.35 (Poor) | 25 ± 2.75 (Good) | 0.001 |

## Discussion

In this study, the knowledge on use of antibiotics only for bacterial infections improved from 11.5% to 82.5%. In the study done by Azevedo et al they have found that the knowledge of correct use of antibiotics for bacterial diseases rather than viral rose from 43% to 76% after the teaching activity (5). In this study, the awareness on when not to use and when to use antibiotics for common infections like respiratory tract infections and gastro enteritis improved from 5.1% to 96.77%. The awareness on use of antibiotics for urinary infection and skin and soft tissue infection improved from 19.6% to 90.38%. The practice of not buying antibiotics from pharmacy, and completing the antibiotic course improved from 20.3% to 91.92%. There is lack of studies conducted which are similar to ours and hence many literatures were not available to compare and correlate.

In our study, out of twelve schools, one school had an excellent performance, while nine schools had a good performance and two schools had a fair performance post intervention. After the intervention, the knowledge on ill-effects of antibiotics improved from 2.5% to 82.5%. A study conducted by Azevedo et al, the knowledge on the risk of resistance to antibiotics by their irrational use rose from 48% to 74% after the teaching activity(5).

During the study we observed that wherever the teachers had a good commitment to get the students educated, the post test results of those students was much better. This indicates that training the school teachers on rational use of antibiotics would help their students to learn on this issue better and help improve the awareness among the school children. We presume that these interventions can be replicated in other schools. Repeated awareness programs should be conducted to promote long lasting effect on knowledge, awareness and practice of rational use of antibiotics.

Studies from countries like Kuwait, Arabic countries, Tanzania, Malaysia, Ethiopia have concluded that the study population have low knowledge and awareness on antibiotic use and antimicrobial resistance (AMR). Many misconceptions prevail on the use of antibiotics for viral infections like flu and all types of fever. For these infections, people demand antibiotics from the doctors and doctors prescribe antibiotics to meet the patient’s expectation. The patients are found to discontinue the antibiotics once they feel symptomatically better. These studies have concluded that dissemination of awareness on rational use of antibiotics is essential to fight against the raising antimicrobial resistance (21–25).

In contradiction to all the above studies, a study from Romania has concluded that the study participants had adequate knowledge of antibiotics and their rational use (19).

There is an urgent need to raise the awareness on antimicrobial resistance and rational use of antibiotics in the wider population. It was observed that even the well-educated adults did not have a clear idea that antibiotics can be used only in bacterial infections. To reach out to the larger population, education through television, radios, YouTube lectures and documentary movies can be used. Educating people using social media like WhatsApp, Facebook, Instagram, LinkedIn, Twitter etc. can also be exploited for this purpose. Many of the other health awareness days like World Diabetes Day, World Tuberculosis Day, World Heart Day, etc. have a wider publicity in the media but the World Antibiotic Awareness Week lacks any kind of attention. Hence, the World Antibiotic Awareness Week should be celebrated more widely.

With the rising incidence of diabetes, immuno-compromised states (Cancer chemotherapy, HIV-AIDs, Immuno-suppressive therapy in transplant patients and auto-immune diseases) across the globe, antibiotics need to be preserved to treat infections in these patients also. Antibiotics are not only used to treat infections but also used prophylactically in the above said immuno-compromised conditions to save lives.

## Conclusion

The knowledge, awareness and practice of rational use of antibiotics is very poor among general public. With raising AMR and to avoid entering to post-antibiotic era, the need of the hour is to create awareness on rational use of antibiotics not only to health care providers but also to the pharmacists and general public. Greater publicity is required on rational use of antibiotics and antimicrobial resistance through all types of media. The World Antibiotic Awareness Week should become a national agenda to raise awareness. Innovative gamified interventions create better and long-lasting awareness on this burning global issue.

## Data Availability

All data produced in the present work are contained in the manuscript

## Acknowledgment

We would like to acknowledge the timely permission given by JSS Mahavidyapeetha for conducting these studies during the World Antimicrobial Awareness week. We also thank the school principals and teachers for their kind cooperation during our study.

## Author Contributions

SMN identified the need of the education, devised the innovative games on awareness and designed the questionnaire and the study. She also got involved in conducting the study, analysis of the report compiled and contributed to prepare the manuscript. SRS and YM prepared the material required for the games, conducted the study in all the schools, compiled the results of the study and helped in manuscript preparation. PK contributed in the statistical analysis of the compiled data.

## Data availability statement

No additional data available.

## Ethical clearance

Name of Ethical Committee: Institutional Ethical Committee, JSS Medical College, JSSAHER.

This study involves human participants but an Ethics Committee(s) or Institutional Board(s) exempted this study because the study did not involve any diagnostic or therapeutic intervention program involving any human subjects. The awareness program was conducted to high school students to educate about rational use of antibiotics and informed consent from each school was taken prior to the scientific awareness program.

